# Effectiveness of a 12-week multicomponent occupational lifestyle intervention to increase physical activity among Japanese teleworkers: a cluster randomised controlled trial (TELEWORK study)

**DOI:** 10.64898/2026.03.23.26349125

**Authors:** Jihoon Kim, Yoshio Nakata, Aya Wada, Natsumi Yoshioka, Kaori Yoshiba, Yuko Kai, Satoru Kanamori, Takahiko Yoshimoto, Rumi Tsukinoki, Wataru Umishio, Tomoko Shiomitsu, Masahiko Gosho

**Affiliations:** Institute of Health and Sport Sciences, University of Tsukuba, Ibaraki 305-8574, Japan; Physical Fitness Research Institute, Meiji Yasuda Life Foundation of Health and Welfare, Tokyo 192-0001, Japan; Graduate School of Public Health, Teikyo University, Tokyo 173-8605, Japan; Department of Hygiene, Public Health and Preventive Medicine, Showa Medical University School of Medicine, Tokyo 142-8555, Japan; Department of Public Health Nursing, Institute of Science Tokyo, Tokyo 113-8519, Japan; Department of Architecture and Building Engineering, School of Environment and Society, Institute of Science Tokyo, Tokyo 152-8552, Japan; School of Nursing, The International University of Kagoshima, Kagoshima 891-0197, Japan; Department of Biostatistics, Institute of Medicine, University of Tsukuba, Tsukuba, Ibaraki 305-8575, Japan

**Keywords:** Occupational health, Health promotion, Work from home, Intervention

## Abstract

**Background:** Teleworking is associated with lifestyle risk factors, such as insufficient physical activity (PA) and increased sedentary time (ST); however, effective interventions tailored to teleworkers are lacking. We aimed to evaluate the effectiveness of a 12-week multicomponent occupational lifestyle intervention on daily step counts among Japanese teleworkers.

**Methods:** This 12-week, two-arm, parallel-group, cluster randomised controlled trial conducted across 12 clusters in six Japanese companies involved 310 teleworkers (mean age 43.0 years; 72.6% men) randomized to the intervention (6 clusters, n=156) or a waitlist control group (6 clusters, n=154). The multicomponent occupational lifestyle intervention included individual (online lectures, feedback, and email messages), physical (posters and a pop-up), and organizational (encouraging messages from an executive) strategies. The primary outcome was the change in daily step counts, measured using an accelerometer over 14 consecutive days at baseline and at week 12. Analyses were based on the intention-to-treat approach using a generalised estimating equation model.

**Findings:** The intervention group showed an adjusted mean change in daily step counts of +219 steps (95% confidence interval [CI] −165 to 604), compared with +188 steps (95% CI −183 to 560) in the control group. The adjusted between-group difference was +55 steps (95% CI −550 to 660; p=0.844). No significant effects on the secondary outcomes (ST, light PA, or moderate-to-vigorous PA) were observed.

**Interpretation:** The multicomponent occupational lifestyle intervention did not significantly increase daily step counts among Japanese teleworkers. Therefore, further studies should be done on tailored interventions for teleworkers.

**Research in context:** *Evidence before this study:* Teleworking has increased globally, particularly following the coronavirus disease pandemic, and has been associated with reduced physical activity and increased sedentary behaviour, both of which are risk factors for cardiovascular disease. Previous studies have also reported that telework environments may contribute to musculoskeletal and other somatic symptoms. Multicomponent interventions in traditional office settings can effectively increase physical activity and reduce sedentary time. These interventions commonly employ strategies at multiple levels of the social–ecological model, including individual approaches (e.g., lectures or incentives), interpersonal approaches (e.g., team-based activities), environmental modifications (e.g., office rearrangements or sit-stand desks), and organisational support (e.g., leadership encouragement). The applicability of such interventions to teleworking populations remains unclear because teleworkers face distinct challenges such as social isolation, blurred work–life boundaries, and heterogeneous home working environments. These contextual differences highlight the need for interventions specifically tailored to teleworkers.

*Added value of this study:* We evaluated the effectiveness of a multicomponent occupational lifestyle intervention specifically designed for teleworkers, a population whose work environment differs substantially from traditional office settings. Our findings provide novel evidence that can inform the development of more targeted strategies to promote physical activity in evolving work environments. This study also provides objective measurements of physical activity using accelerometers, enabling detailed evaluation of step counts, sedentary time, and different activity intensities among teleworkers. Additionally, we used daily diaries to distinguish activity patterns across workdays, weekends, teleworking days, and commuting days, providing a nuanced understanding of behavioural patterns in remote work contexts.

*Implications of all the available evidence:* Our findings indicate that low-cost, remotely delivered multicomponent interventions may be insufficient to produce meaningful behavioural change among teleworkers. Similar strategies, including individual, physical, and organisational components, have been effective in traditional workplace interventions; however, their implementation in teleworking environments may not adequately address the specific challenges faced by remote workers. In particular, the lack of strong sociocultural support mechanisms, such as team-based step competitions or workplace champions, may limit engagement and reduce the effectiveness of such programmes. Therefore, further investigation is needed to explore more diverse and targeted intervention strategies, identify the specific needs and behavioural patterns of teleworkers, and apply more precise eligibility criteria to better address heterogeneity within teleworking populations.

## Introduction

The global work environment has undergone a marked shift with the widespread adoption of teleworking, particularly following the coronavirus disease (COVID)-19 pandemic.^1^ In Japan, the proportion of workers engaging in telework increased from 14.8% in the year preceding the COVID-19 pandemic to 23% immediately afterwards and has since stabilised at 24.6% as of 2024.^2^ While teleworking offers benefits such as reduced commuting time and greater flexibility in daily schedules, it is also associated with increased sedentary time (ST) and decreased physical activity (PA),^3,4^ which are established risk factors for cardiovascular disease.^5,6^ Moreover, telework environments are often ergonomically inadequate compared to office settings, which may lead to musculoskeletal and somatic symptoms.^7,8^ Targeted interventions to address these health challenges are critical for teleworkers.

Numerous well-designed interventions have been effective at increasing PA and reducing ST in traditional office settings.^9–12^ A common characteristic of these interventions is their multicomponent design based on the social–ecological model, targeting not only individuals (e.g. lectures and incentives) but also including interpersonal (e.g. team-based activities), environmental (e.g. office rearrangements and installation of sit-stand desks), and organisational (e.g. leadership support) strategies.^9–13^ However, their efficiency in teleworkers, with their unique context marked by social isolation,^14^ blurred work–life boundaries, and variable work environments,^15^ is unknown. Thus, developing interventions tailored to teleworking contexts is necessary.

To address this gap, in this TELEWORK study, we evaluated the effectiveness of a multicomponent occupational lifestyle intervention for improving PA and related health outcomes among Japanese teleworkers. The intervention focused on three key areas: increasing PA, enhancing musculoskeletal health, and optimising the remote work environment by considering the unique working conditions of teleworkers compared to those of office workers.^16^ A cluster-randomised controlled trial design was employed to minimise the risk of contamination, as the social components of the intervention could have led to information sharing among individuals within the same workplace.^17,18^ Through this study, we aim to generate evidence to inform effective PA promotion in future occupational health strategies for teleworkers.

## Methods

### Trial design

We conducted a 12-week, two-arm, parallel-group, cluster-randomised controlled trial to evaluate the effectiveness of a multicomponent occupational lifestyle intervention for increasing PA among Japanese teleworkers. This trial was conducted in Japan from March 2024 to March 2025 and adheres to the CONSORT 2025 statement^19^ and its extension for cluster randomised trials.^20^

### Participants, setting, and clusters

This study was conducted among workers employed in companies in the Tokyo metropolitan area of Japan. We initially approached 16 companies, of which six agreed to participate in the trial following initial screening and institutional approval, yielding 12 distinct clusters for randomisation. The participating companies represented a range of industries, including information technology, trading, and automobile manufacturing. Companies were identified and invited via researchers’ academic networks, using a snowball recruitment approach.

Clusters were defined as organizational units within each participating company, such as departments, teams, or office branches, to reflect existing workplace structures and minimise contamination between participants. Within each participating company, recruitment was coordinated through human resource departments and internal email announcements. Each participant received the study information and volunteered to participate. Based on company and organizational size, clusters typically comprised 20–100 workers.^16^

We included participants (1) aged 18–65 years and (2) working from home at least 1 day per week. We excluded individuals who were (1) prohibited from exercising by a doctor, (2) planning long-term business trips or retirement during the study period, (3) pregnant or possibly pregnant, and (4) judged to be inappropriate for inclusion by the research director (e.g. not agreeing to return the measuring instruments). Teleworking frequency varied depending on departmental policies and personal status.

### Randomization and masking

Clusters were randomly assigned in a 1:1 ratio to either the intervention or waitlist control group using stratified block randomisation by cluster size (<20 or ≥20 participants). An independent statistician (M.G.) generated the allocation sequence with a fixed block size of two, using a computerised random number table. The allocation list was provided to an independent research coordinator affiliated with the Meiji Yasuda Life Foundation of Health and Welfare as clusters were enrolled. Allocation concealment was not fully maintained, and the participants, facilitators, and outcome assessors were aware of the group allocation. However, PA was objectively measured using a triaxial accelerometer, which did not display the data to the participants.

### Interventions

The 12-week multicomponent occupational lifestyle intervention was theoretically grounded on the social–ecological model^21^ and delivered exclusively through a remote system comprising two key platforms: (1) a dedicated website that hosted all educational lectures and resources, and (2) weekly email messages used for participant support. This theoretical model guided the program’s structure, which utilised individual, sociocultural, physical, and organizational strategies to ensure comprehensive support for behavioural changes among teleworkers. The intervention had three principal objectives: increasing PA, enhancing musculoskeletal health, and optimising the remote work environment. Implementation was successfully supported by a designated workplace coordinator in each cluster, which enabled minor site-specific adaptations.

### Individual strategies

Individual strategies were delivered through three primary sources: online lectures, feedback, and emails. First, a dedicated website allowed participants to access online lectures at their convenience. The content was structured according to the three intervention objectives. The PA promotion section featured three theoretical lectures (4–5 min each) presented by a researcher (Y.N.) that covered the definitions of PA and ST, the health impact of physical inactivity, and evidence of +10 min of PA per day.^22–24^ Moreover, five specific exercise videos (2–5.5 minutes each) were provided, focusing on locomotor training and 3-min stretching routines for workers (produced by Science Tokyo and Nomura Real Estate Group).^25^ The website also hosted brief educational videos related to two other objectives: musculoskeletal health and remote work environment optimisation. The detailed content of these non-PA educational materials is available in the study protocol.^16^

Participants in the intervention group received feedback in the form of an occupational report addressing all three topics. The PA section of this report provided participants with detailed feedback based on their objectively measured baseline results, helping them understand their current PA status before the intervention. This included daily step counts; daily time spent in light-intensity PA (LPA), moderate-intensity PA, vigorous-intensity PA, and moderate-to-vigorous intensity PA (MVPA); total weekly MVPA; and daily ST.

A key feature of this feedback was that all PA data were presented in two formats: (1) as an overall daily average and (2) as a specific average for teleworking days only. This allowed participants to identify specific behavioural patterns associated with telework settings. To help participants interpret these results, the report included explicit descriptions of each activity intensity level and the results of their PA against the current Japanese and World Health Organization PA guidelines.^26,27^ This contextual information enabled participants to assess the adequacy of their own PA levels. The report also included brief feedback on musculoskeletal health (i.e. back pain) and the telework environment (i.e. environmental sensor measurements). Full details regarding the non-PA components of the feedback report are provided in the study protocol.^16^

The research team sent emails on practical tips to the workplace coordinator over the 12 weeks on the three intervention objectives twice weekly (Tuesdays and Fridays). These emails were then disseminated to the participants. To improve adherence, coordinators were encouraged to ask their supervisors to lead the most recently distributed exercises in group settings.

### Sociocultural environmental strategies

The sociocultural environmental strategy was designed to foster a supportive atmosphere and promote team-building among participants. The plan was an individual-and team-based step-count competition. Individual competition was intended to increase personal engagement with the programme, whereas team-based competition was aimed at enhancing social support and cohesion within clusters.

### Physical environmental strategies

The physical environmental strategy consisted of two primary materials provided to the intervention group: PA promotion posters and tabletop pop-ups. PA-focused posters were made accessible either on the office noticeboard or a dedicated intervention website. The topics focused on the health risks of prolonged sitting, the benefits of standing and walking, and the importance of goal-setting for increasing daily PA. The tabletop pop-up was designed for participants’ work desks and provided illustrated instructions for a specific stretch aimed at managing back pain.

### Organizational strategies

The organizational strategy consisted of encouraging messages from the upper management to participants in the intervention group. The content of these messages was determined by each company’s management and emphasised the importance of participation and provided specific encouragement. These encouraging messages were delivered via email (described in the individual strategies section), whereby the research team coordinated with the workplace coordinators for dissemination.

### Control group

The control group served as a waitlist group and received no intervention during the 12-week intervention period, but they were instructed to maintain their usual work and lifestyle habits. After the 12-week follow-up data collection, the intervention program was offered to the control group clusters if their companies wished to receive it.

### Primary and secondary outcomes

The primary outcome was the change in daily step count over the 12-week intervention period. Secondary outcomes included changes in ST, LPA, and MVPA, assessed over the same period. PA was objectively measured using a triaxial accelerometer (Active style Pro HJA-750C; Omron Healthcare, Kyoto, Japan), a validated device for estimating step counts and activity intensity in free-living adults, employing a proprietary algorithm to calculate daily steps and classify activity intensity (expressed as metabolic equivalents).^28,29^

Participants were instructed to wear the device on their waist for 14 consecutive days at baseline (week 0) and follow-up (week 12). Participants were asked to remove the device only when sleeping, bathing, swimming, or engaging in contact sports. Accelerometer data were considered valid when the device was worn for ≥10 h per day for ≥3 days.^30^ Participants also completed a daily diary to record contextual information, including working hours and work type (teleworking or commuting workdays).

Moreover, baseline demographic (age, sex), socioeconomic (job position, years of service, education level, living status, marital status, household income), lifestyle (sleep time, PA activity habits, smoking status, alcohol use, dietary habits, weekly working hours, and weekly teleworking frequency), and clinical characteristics (self-reported height and weight, body mass index) were collected using a self-reported electronic questionnaire. .

### Other secondary outcomes

We also collected data addressing two additional intervention objectives: musculoskeletal health and optimisation of the remote work environment. Details of these outcomes, including the specific measurement instruments and assessment procedures, are described in the study protocol.^16^ Given our focus on PA as an outcome, detailed analyses and results concerning musculoskeletal and remote work environment outcomes will be reported separately in subsequent publications. A post-intervention assessment of the program was also conducted.

### Safety and adverse events

Designated workplace coordinators at each participating company monitored adverse events. Participants were instructed to report any study-related injuries, persistent pain, or health concerns to their respective workplace coordinators, who then relayed the information to the research team for appropriate medical evaluation and response.

### Sample size calculation

The required sample size was predetermined, as shown in the study protocol.^16^ The sample size was calculated to detect a significant difference in daily step count between the two treatment groups over the 12-week intervention. Based on a precious study, the effect size of the intervention was estimated at Cohen’s d = 0.33,^16^ corresponding to an estimated difference between groups of 800 steps considered clinically meaningful,^31^ assuming a standard deviation (SD) of 2,400 steps. The calculation was performed using a two-sided test with a type I error rate of 0.05 and a statistical power of 0.80. Based on these parameters, the required sample size for an individually randomised trial was 146 participants per group, totalling 292 participants.

The sample size was inflated to account for the non-independence of the data inherent in the cluster randomized controlled trial design and to accommodate within-cluster correlation. Based on the assumed average cluster size of 20 participants and an intraclass correlation coefficient of 0.02, the adjusted required calculated sample size was 403 participants. Finally, anticipating a 20% dropout rate (i.e. 80% completion rate), the final recruitment target was set at 500 participants.

### Statistical analysis

Continuous variables are presented as mean (SD) or median (interquartile range), depending on the data distribution, while categorical variables are expressed as numbers and percentages (n, %). All statistical analyses were based on the full analysis set (FAS), defined according to the intention-to-treat (ITT) principle. The FAS included all randomised participants, regardless of protocol adherence or withdrawal from the study.

Missing data were handled using multiple imputation by chained equations with 500 imputed datasets. The imputation model included all outcome variables, baseline covariates, and auxiliary variables predictive of missing data. The missing-at-random assumption was considered plausible, given that missingness was associated with observed baseline characteristics. Estimates from each imputed dataset were pooled using Rubin’s rules, with adjusted degrees of freedom for small samples.^32^

The primary (change in daily step count) and secondary outcomes (changes in ST, LPA, and MVPA) were analysed using GEE with an exchangeable correlation structure, accounting for clustering at the worksite level in accordance with the study protocol.^16^ The model included group assignment (intervention and control), time (baseline and week 12), and group × time interaction. Accelerometer wear time was included as a covariate. Given the small number of clusters (n=12), a bias-corrected robust variance estimation (CR2) was applied, as recommended for randomised cluster trials with few clusters.^33^ Specifically, a CR2 estimator with Satterthwaite degrees of freedom was used.^34^

Sensitivity analysis was conducted using the per-protocol set (PPS). PPS was defined as participants who met all the following criteria: >65 years of age, engaged in telework at least 1 day per week before and after the intervention, and received the minimum intervention (i.e. confirmed viewing of intervention emails). In another sensitivity analysis, GEE without multiple imputations was applied to the outcomes, assuming that data were missing completely at random. Subdomain analysis was conducted using the same GEE method with multiple inputs. This analysis explored the difference in the intervention effect across work-, weekend-, teleworking-, and commuting days. All statistical tests were two-sided, with significance set at p<0.05. All analyses were performed using R version 4.4.1 with the mouse, geepack, clubSandwich, and lme4 packages.

### Ethics and registration

This trial was approved by the Meiji Yasuda Life Foundation of Health and Welfare (approval number: 2023-0002) and registered in the University Hospital Medical Information Network Clinical Trials Registry (UMIN000053861) on 15 March 2025, prior to participant enrolment. The full trial protocol has been published previously.^16^

### Results Participant flow

We assessed 370 individuals across 12 clusters from six companies for eligibility. Of these, 60 individuals were excluded for the following reasons: 39 declined to provide consent, 8 did not provide baseline data, and 13 were research collaborators who were deemed inappropriate for inclusion by the research director. Finally, 310 participants across 12 clusters were randomised: 6 clusters (156 participants) to the intervention group and 6 clusters (154 participants) to the control group (median cluster size 28 [18–33] and 25 [18–33] participants, respectively). During the week 12 follow-up period, 19 participants (12.2%) in the intervention group and 29 participants (18.8%) in the control group were lost to follow-up (did not provide outcome data for week 12).

Of the 156 participants assigned to the intervention group, 137 (87.8%) completed the postintervention assessment. Among these participants, adherence was highest for checking emails (n=121, 88.3%), followed by feedback (n=105, 76.6%), receiving encouraging messages (n=93, 67.9%), online lectures about PA (n=75, 54.7%), checking PA promotion posters (n=64, 46.7%), and watching online lectures about exercise (n=63, 46.0%).

Primary statistical analyses were conducted using FAS, which included all 310 randomised participants according to the ITT principle. The flow of participants and clusters during the trial is shown in Figure 1.

**Figure 1.**
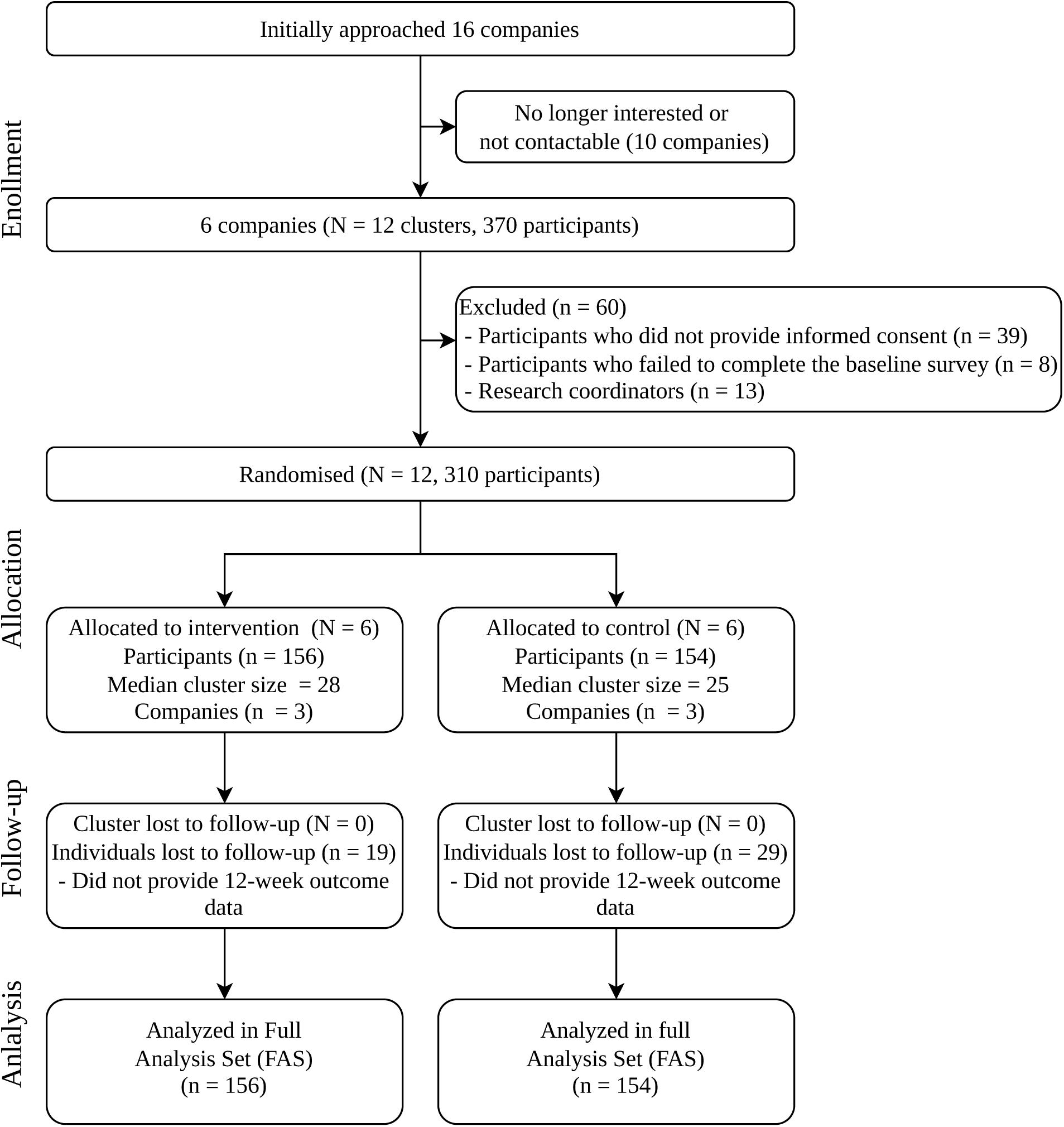
Flow diagram.

### Baseline characteristics

The intervention and control groups were similar at baseline across all measured characteristics. At the individual level, the mean age was 42.4 (SD 12.0) years and 43.7 (SD 11.4) years in the intervention and control groups, respectively, and most participants were men (76.9% vs. 68.2%, respectively). The proportions of participants engaging in PA for ≥60 min/day were 43.6% and 40.3% in the intervention and control groups, respectively. Telework frequency (≥1 day/week) was also comparable (71.2% vs. 75.3%). Of the 310 participants, 283 (91.3%) provided valid baseline accelerometer data. Mean daily step count was 7,036 (SD 2,803) in the intervention group and 6,941 (SD 2,971) in the control group (Table 1).

**Table 1.** Baseline characteristics.

| Characteristic | Intervention | Control | Total |
| --- | --- | --- | --- |
| **Cluster-level** |  |  |  |
| Number of companies | 3 | 3 | 6 |
| Number of clusters | 6 | 6 | 12 |
| Cluster size <20, n (%) | 1 (17) | 2 (33) | 3 (25) |
| Cluster size ≥20, n (%) | 5 (83) | 4 (67) | 9 (75) |
| Cluster size, median (IQR) | 28 (18–33) | 25 (18–33) | 26 (19–32) |
| **Individual-level** |  |  |  |
| Number of participants, n | 156 | 154 | 310 |
| Age, years, mean (SD) | 42.4 (12.0) | 43.7 (11.4) | 43.0 (11.7) |
| Sex, men, n (%) | 120 (76.9) | 105 (68.2) | 225 (72.6) |
| Height, cm, mean (SD) | 169.1 (7.7) | 168.1 (7.8) | 168.6 (7.7) |
| Weight, kg, mean (SD) | 67.9 (13.2) | 64.8 (12.6) | 66.4 (13.0) |
| Body mass index, kg/m<sup>2</sup>, mean (SD) | 23.6 (3.7) | 22.8 (3.7) | 23.2 (3.7) |
| Lifestyle status |  |  |  |
| Sleep time, hours/day, mean (SD) | 6.2 (0.9) | 6.1 (0.8) | 6.2 (0.9) |
| Physical activity ≥ 60 min/day, n (%) | 68 (43.6) | 62 (40.3) | 130 (41.9) |
| Current smoker, n (%) | 19 (12.2) | 24 (15.6) | 43 (13.9) |
| Alcohol use ≥1 day/week, n (%) | 80 (51.3) | 78 (50.6) | 158 (51.0) |
| Vegetable or fruit consumption every day, n (%) | 74 (47.4) | 84 (54.5) | 158 (51.0) |
| **Socioeconomic status** |  |  |  |
| Job position, n (%) |  |  |  |
| Non-regular worker | 8 (5.1) | 7 (4.5) | 15 (4.8) |
| Regular worker | 100 (64.1) | 110 (71.4) | 210 (67.7) |
| Manager | 48 (30.8) | 37 (24.0) | 85 (27.4) |
| Years of service, years, median (IQR)<sup>†</sup> | 14 (5–25) | 15 (4–24) | 15 (5–24) |
| Four-year college graduate, n (%) | 123 (78.8) | 124 (80.5) | 247 (79.7) |
| Living with others, n (%) | 111 (71.2) | 111 (72.1) | 222 (71.6) |
| Currently married, n (%) | 95 (60.9) | 93 (60.4) | 188 (60.6) |
| Household income ≥ 7 million JPY, n (%) | 93 (59.6) | 88 (57.1) | 181 (58.4) |
| Telework frequency ≥ 1 day/week, n (%) | 111 (71.2) | 116 (75.3) | 227 (73.2) |
| Work hours ≥ 40 hours/week, n (%) | 131 (84.0) | 116 (75.3) | 247 (79.7) |
| Accelerometer-measured physical activity n = 283‡ |  |  |  |
| Daily steps, steps/day, mean (SD)‡ | 7,036 (2,803) | 6,941 (2,971) | 6,989 (2,883) |
| Light-intensity physical activity, min/day, mean (SD)‡ | 185.8 (54.6) | 177.1 (67.2) | 181.4 (61.3) |
| Moderate to vigorous intensity physical activity, min/day, mean (SD)‡ | 54.2 (24.2) | 54.1 (24.9) | 54.2 (24.5) |
| Sedentary time, min/day, mean (SD)‡ | 638.0 (106.8) | 633.4 (108.1) | 635.7 (107.3) |
| Wear time, min/day, mean (SD)‡ | 878.0 (112.2) | 864.5 (110.9) | 871.3 (111.6) |
| Valid days, median (IQR)‡ | 12 (9–13) | 11 (8–13) | 11 (9–13) |

Data are presented as the mean (SD), median (Interquartile range [IQR]), or n (%).
† Median (IQR).
‡ Computed for participants with valid accelerometer data ( $\geq 10$ h/day on $\geq 3$ days). Baseline validation data were available for 142 participants in the intervention group and 141 participants in the control group. Participants with fewer valid days were retained in the full analysis set but were treated as missing for the accelerometer-measured physical activity variables. IQR, interquartile range; SD, standard deviation.

### Protocol deviation

The protocol included individual- and team-based step-count competitions; however, these components were not implemented as planned because some companies already had similar programs and others lacked an environment to support competition.

### Effects on primary and secondary outcomes

Regarding the primary outcome, the intervention group showed a high mean change of daily steps compared to the control group [+219 (SD 2,451) vs. +188 steps (SD 2,581)]. The adjusted mean difference between the groups was +55 steps (95% CI, −550 to 660; p=0.844), which was not significant. Similarly, no significant intervention effects were observed for any secondary outcome. There were no significant differences between the groups in the adjusted mean change for ST (−3.1 min/day; 95% CI, −25.9 to 19.7; p=0.768), LPA (−2.5 min/day; 95% CI, −15.8 to 10.7; p=0.679), or MVPA (+1.4 min/day; 95% CI, −3.1 to 5.9; p=0.501) (Table 2).

**Table 2.**
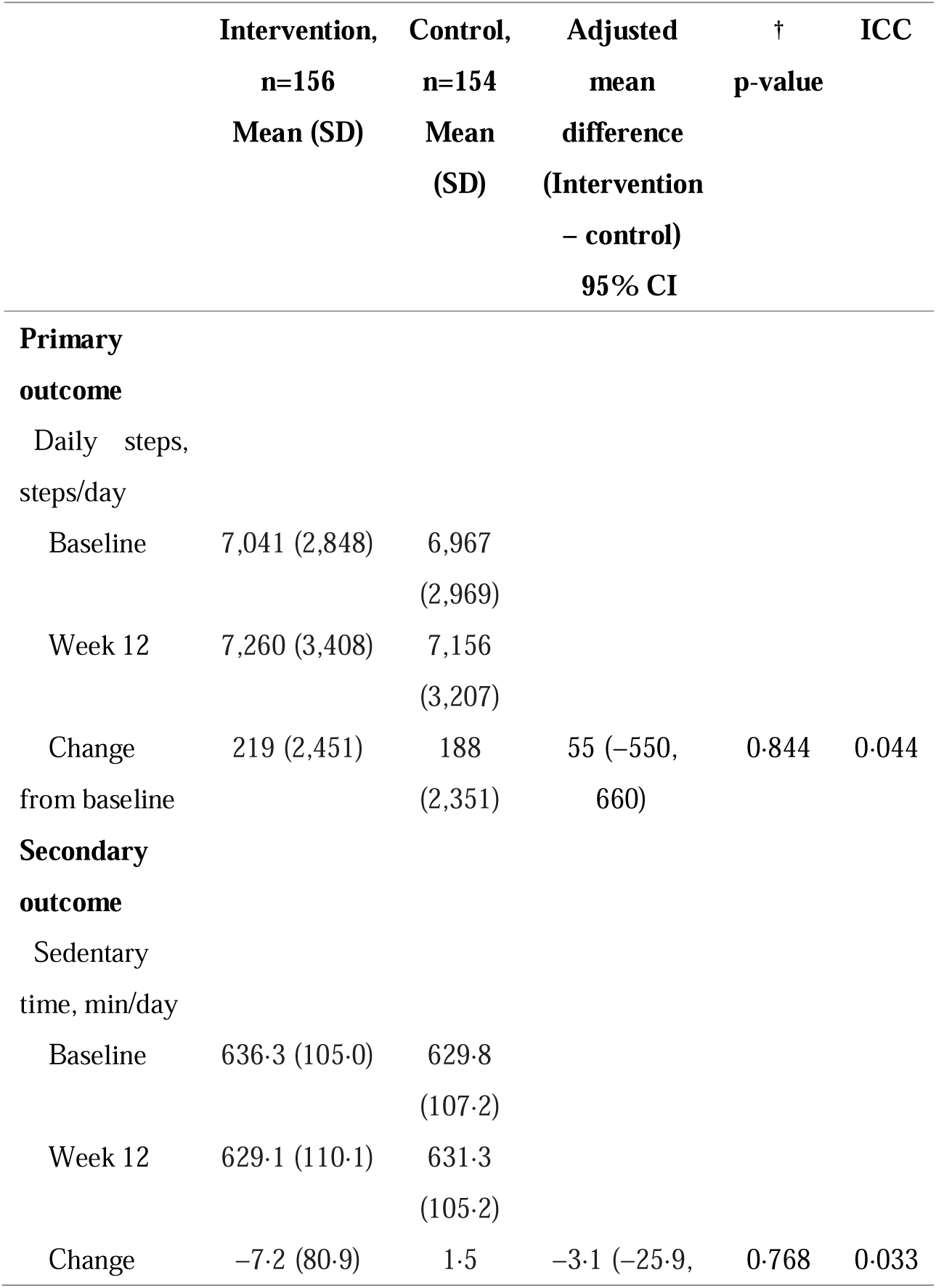
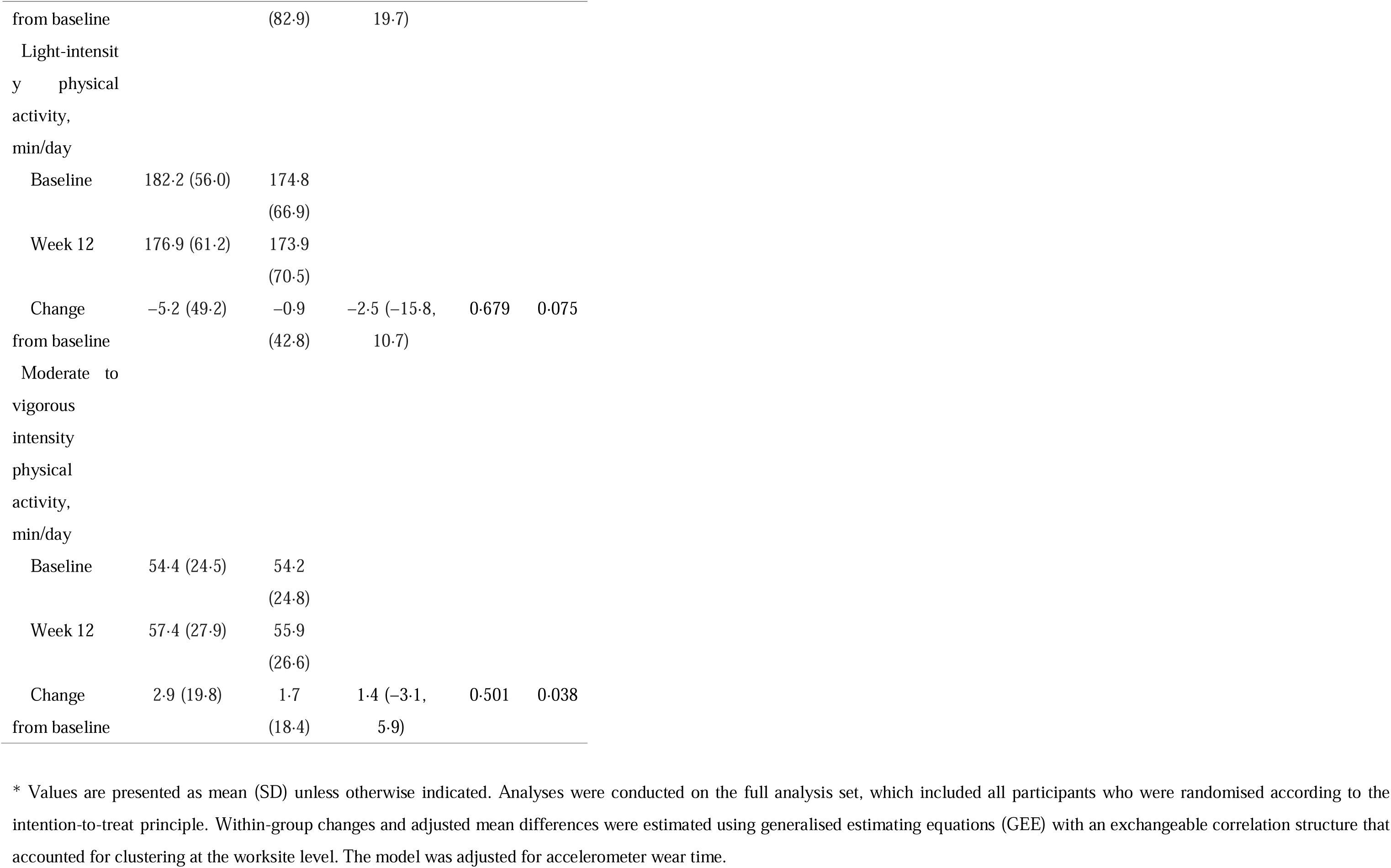
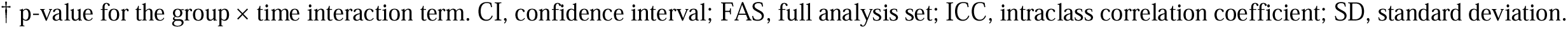
Changes in physical activity outcomes from baseline to week 12 in FAS.

|  | **Intervention,**<br>**n=156**<br>**Mean (SD)** | **Control,**<br>**n=154**<br>**Mean**<br>**(SD)** | **Adjusted**<br>**mean**<br>**difference**<br>**(Intervention**<br>**– control)**<br>**95% CI** | **†**<br>**p-value** | **ICC** |
| --- | --- | --- | --- | --- | --- |
| **Primary** |  |  |  |  |  |
| **outcome** |  |  |  |  |  |
| Daily steps,<br>steps/day |  |  |  |  |  |
| Baseline | 7,041 (2,848) | 6,967<br>(2,969) |  |  |  |
| Week 12 | 7,260 (3,408) | 7,156<br>(3,207) |  |  |  |
| Change<br>from baseline | 219 (2,451) | 188<br>(2,351) | 55 (–550,<br>660) | 0.844 | 0.044 |
| **Secondary** |  |  |  |  |  |
| **outcome** |  |  |  |  |  |
| Sedentary<br>time, min/day |  |  |  |  |  |
| Baseline | 636.3 (105.0) | 629.8<br>(107.2) |  |  |  |
| Week 12 | 629.1 (110.1) | 631.3<br>(105.2) |  |  |  |
| Change | –7.2 (80.9) | 1.5 | –3.1 (–25.9, | 0.768 | 0.033 |
| from baseline |  | (82.9) | 19.7) |  |  |
| Light-intensit |  |  |  |  |  |
| y physical |  |  |  |  |  |
| activity, |  |  |  |  |  |
| min/day |  |  |  |  |  |
| Baseline | 182.2 (56.0) | 174.8 |  |  |  |
|  |  | (66.9) |  |  |  |
| Week 12 | 176.9 (61.2) | 173.9 |  |  |  |
|  |  | (70.5) |  |  |  |
| Change | -5.2 (49.2) | -0.9 | -2.5 (-15.8, | 0.679 | 0.075 |
| from baseline |  | (42.8) | 10.7) |  |  |
| Moderate to |  |  |  |  |  |
| vigorous |  |  |  |  |  |
| intensity |  |  |  |  |  |
| physical |  |  |  |  |  |
| activity, |  |  |  |  |  |
| min/day |  |  |  |  |  |
| Baseline | 54.4 (24.5) | 54.2 |  |  |  |
|  |  | (24.8) |  |  |  |
| Week 12 | 57.4 (27.9) | 55.9 |  |  |  |
|  |  | (26.6) |  |  |  |
| Change | 2.9 (19.8) | 1.7 | 1.4 (-3.1, | 0.501 | 0.038 |
| from baseline |  | (18.4) | 5.9) |  |  |

\* Values are presented as mean (SD) unless otherwise indicated. Analyses were conducted on the full analysis set, which included all participants who were randomised according to the intention-to-treat principle. Within-group changes and adjusted mean differences were estimated using generalised estimating equations (GEE) with an exchangeable correlation structure that accounted for clustering at the worksite level. The model was adjusted for accelerometer wear time.
† p-value for the group × time interaction term. CI, confidence interval; FAS, full analysis set; ICC, intraclass correlation coefficient; SD, standard deviation.

### Sensitivity analysis

Sensitivity analysis was conducted using PPS. This analysis included 174 participants who met all the per-protocol criteria (84 in the intervention group and 90 in the control group). Regarding the primary outcome, the intervention group showed a mean change of +152 steps (SD 2,658), while the control group showed a change of −321 steps (SD 2,294). The adjusted mean difference between the groups was +279 steps (95% CI, −737 to 1,296; p=0.542), which was not significant. Similarly, no significant intervention effects were observed for any secondary outcomes (Table 3).

**Table 3.**
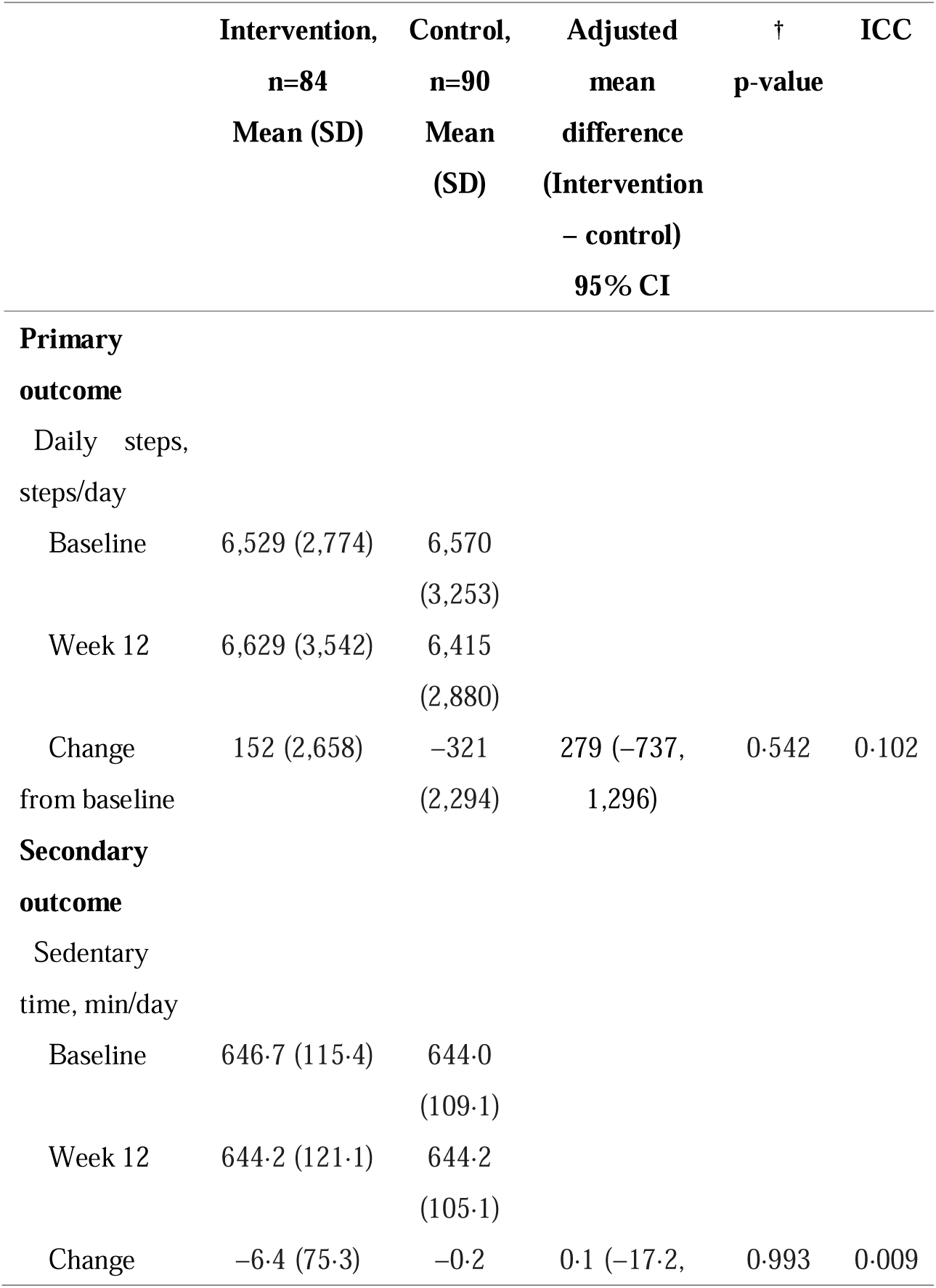
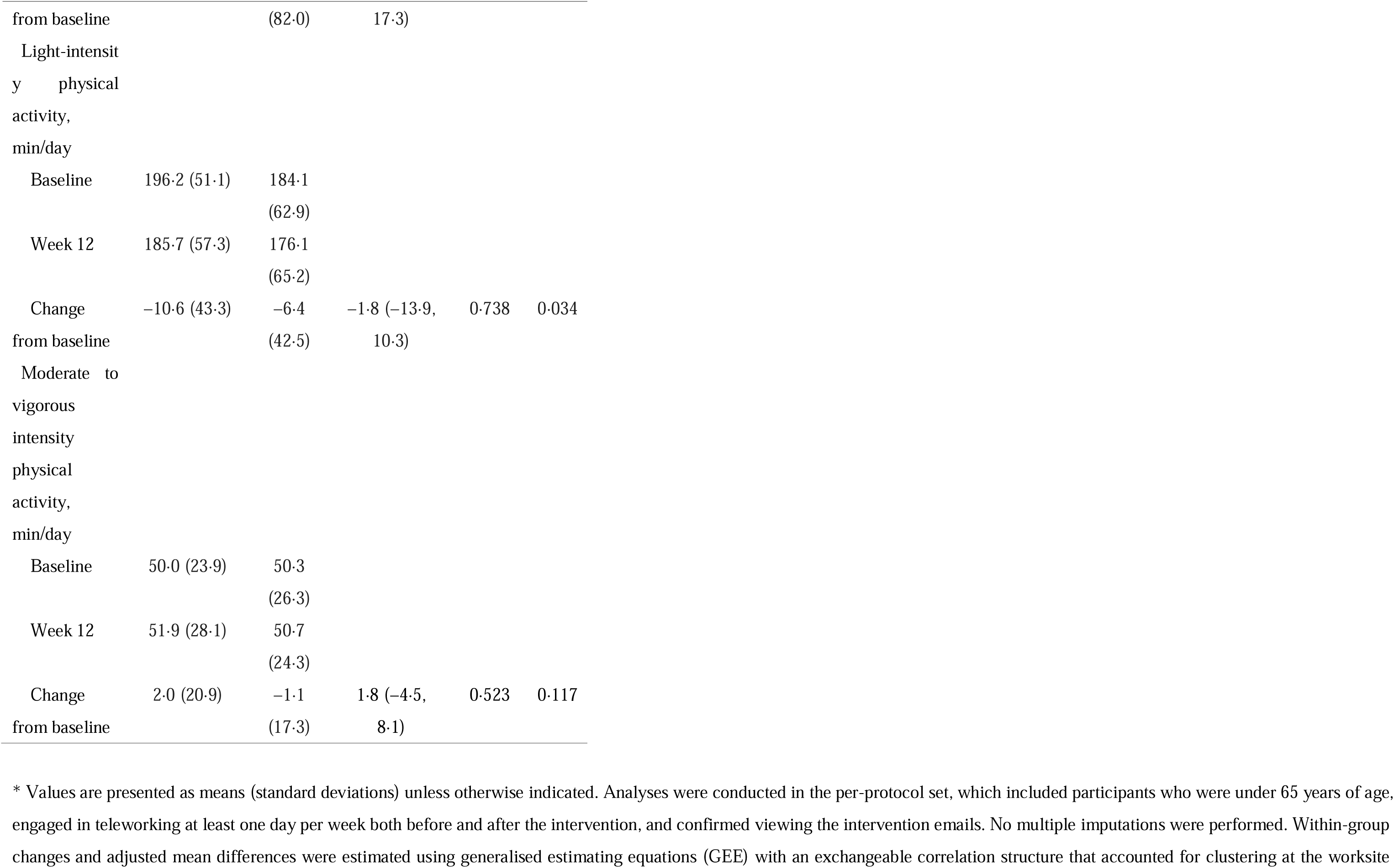
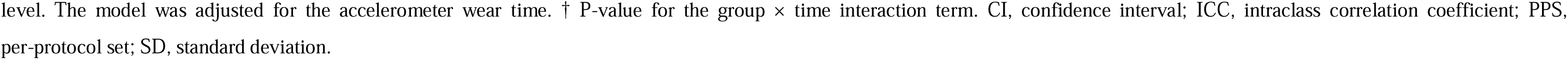
Changes in physical activity outcomes from baseline to week 12 in PPS.

The results of the GEE analysis without multiple imputations were provided in the Supplementary Material (Supplementary Table 1). These results were similar to those of the GEE analysis using multiple imputation.

### Subdomain analysis

Finally, planned subdomain analyses were conducted to explore the difference in the intervention effect by day type (Table 4). Regarding workdays, the intervention group showed a mean change of +100 steps (SD 2,097), while the control group showed a change of +81 steps (SD 1,886). The adjusted mean difference was +34 steps (95% CI, −411 to 479; p=0.868). On weekends, the intervention group showed a change of +614 steps (SD 5,066), whereas the control group showed a decrease of 142 steps (SD 5,145); the adjusted mean difference was +504 steps (95% CI, −1,222 to 2231; p=0.530). For teleworking days, the intervention group showed a change of 188 steps (SD 2,814) compared to 12 steps (SD 2,272) in the control group, resulting in an adjusted mean difference of +178 steps (95% CI, −786 to 1,142; p=0.689). Finally, on commuting days, the intervention and control groups showed a change of −489 (SD 2,081) and −78 steps (SD 1,765), respectively. The adjusted mean difference was −396 steps (95% CI, −966 to 175; p=0.153). Regarding the secondary outcomes (ST, LPA, and MVPA), no significant intervention effects were observed in any of the subdomains (Table 4).

**Table 4.**
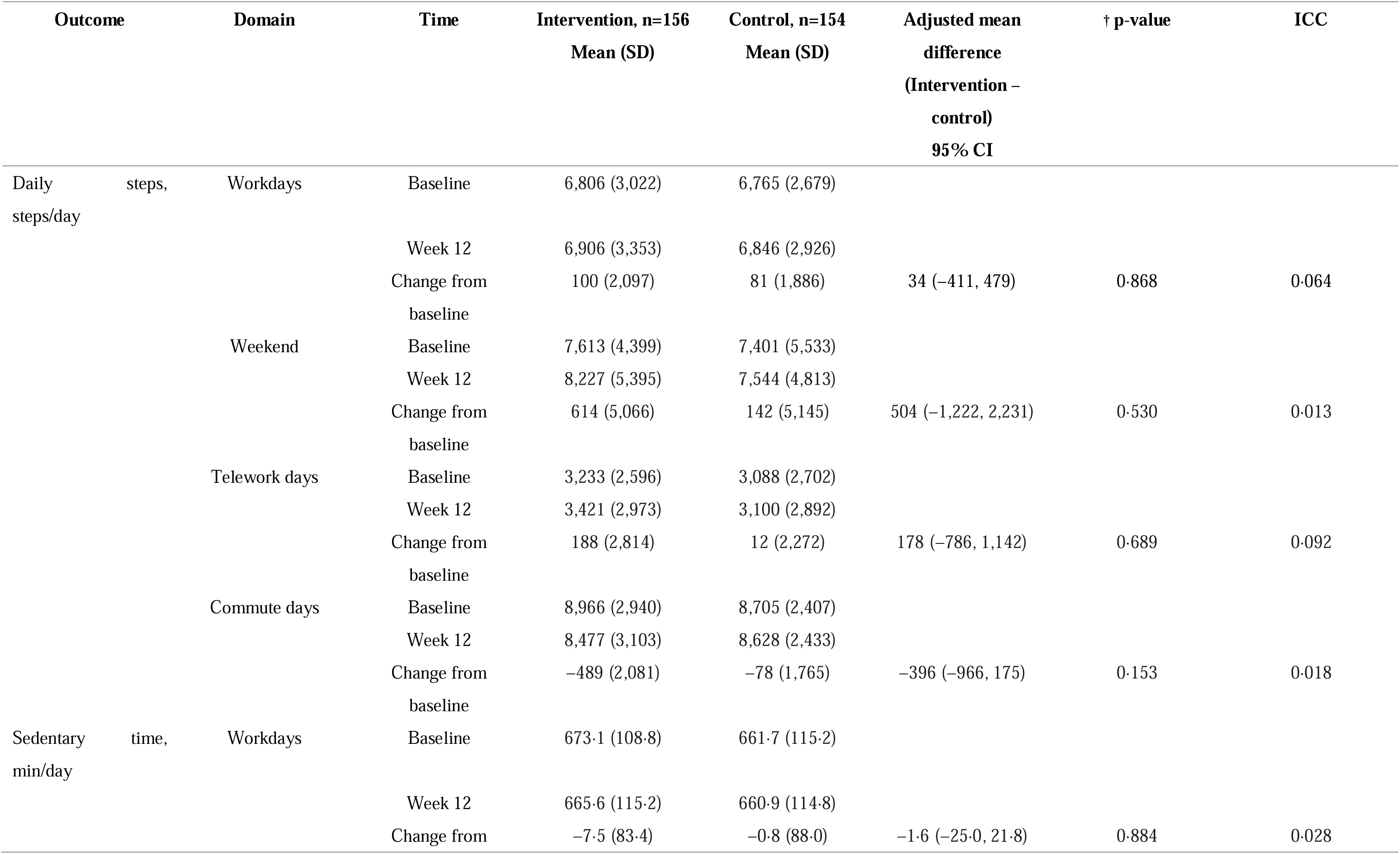
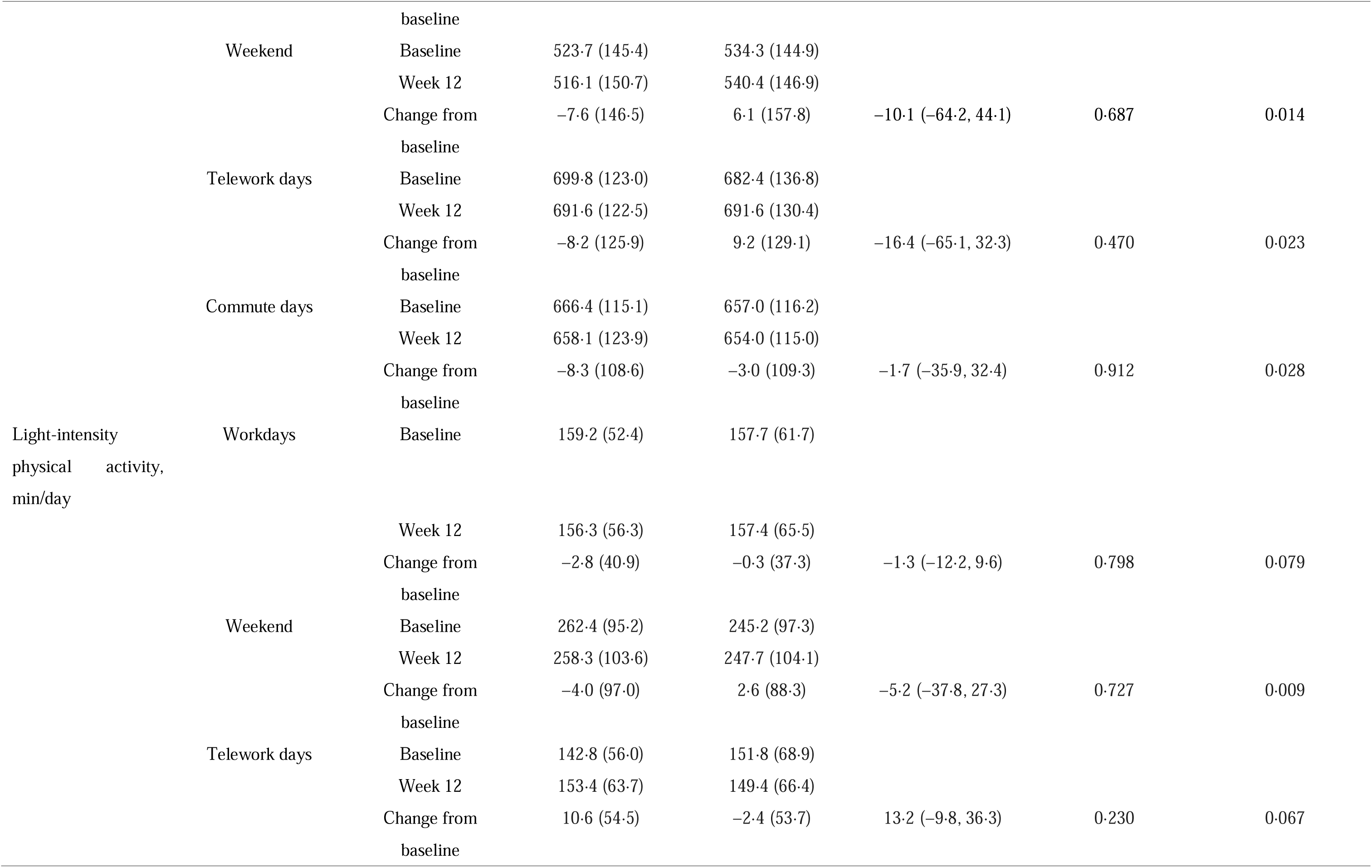
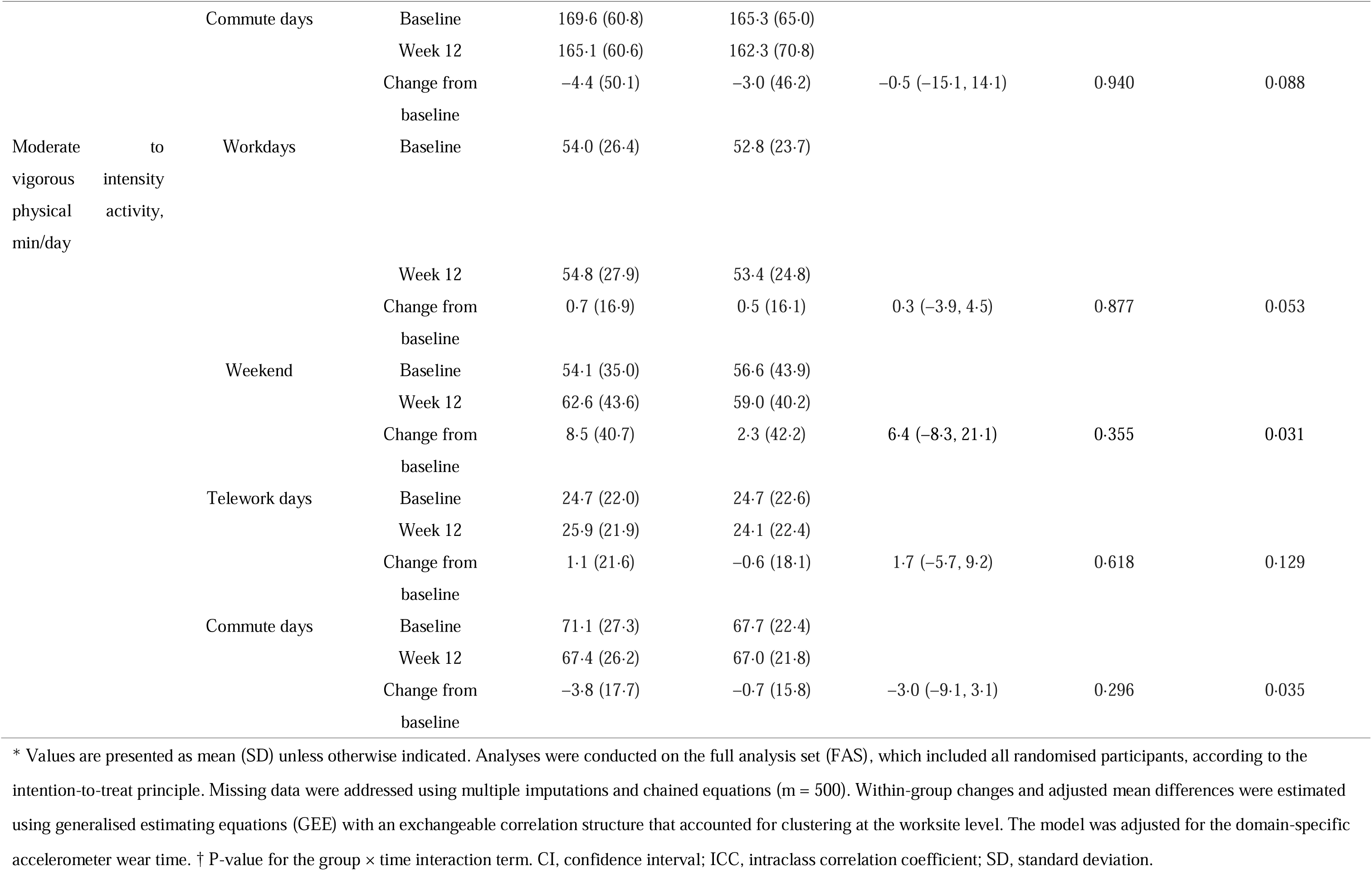
Subdomain analyses of changes in physical activity outcomes from baseline to week 12.

### Adverse events

No serious adverse events or unintended effects attributable to the intervention were reported in either the intervention or control group during the 12-week study period.

## Discussion

This study evaluated the effectiveness of a multicomponent occupational lifestyle intervention for Japanese teleworkers. We found a non-significant increase in the daily step counts in the interventional group. We observed no significant intervention effects on any of the secondary outcomes. Sensitivity analysis also confirmed a non-significant result, as did the exploratory subdomain analyses, which found no significant intervention effects across the different work types (workdays, weekends, commuting workdays, and teleworking workdays).

Our intervention was theoretically grounded in the social–ecological model and included multicomponent interventions that have been reported to be effective in other occupational cluster-randomized controlled trials.^9–13,35^ However, the consistent null finding across all analyses suggests that the implementation or design of these strategies was insufficient to drive behavioural change in the teleworker’s specific context. We presume several critical factors explain this phenomenon.

First, the intervention may lack key components to drive substantial behavioural changes. Specifically, our program did not provide financial incentives, height-adjustable workstations, or wearable devices with real-time feedback,^10–13,36^ which are highly effective in promoting PA. However, these methods incur high adoption costs and are not feasible for most companies. In contrast, our intervention prioritised low-cost, more feasible strategies that could be delivered remotely without placing a financial burden on the participating companies. This is considered the primary reason for the null finding.

Second, a team-based step competition was not implemented. This was planned as the core sociocultural environmental strategy to foster a supportive atmosphere and promote team building. ’Social isolation’ is a core challenge for teleworkers.^14^ Previous studies have shown that intervention components, such as step competitions or the presence of ’workplace champions’ who encourage participation, create a supportive atmosphere and promote team-building.^11–13,37^ In this study, some companies already had similar competition programs, while others had difficulty establishing the necessary environment. Consequently, our program failed to provide this essential sociocultural support and address the isolation teleworkers face, which was likely insufficient to drive behavioural change.

Third, the intervention may not have been optimally targeted for teleworkers. This program was closer to adapted traditional office strategies delivered remotely rather than one specifically designed for the unique context of teleworkers. Consistent with previous studies targeting office workers,^38,39^ future ones should first identify the status and needs of teleworkers to configure appropriate intervention programs. Furthermore, the intervention did not focus exclusively on PA. The program was designed to address three objectives: PA, musculoskeletal health, and the work environment. This diffusion of focus may have diluted the ’dose’ of PA-specific content, weakening the intervention’s potential impact.

Finally, low adherence to specific educational components of the intervention may have contributed to the null finding. Engaging with emails (88.3%) and feedback (76.6%) was high; however, adherence to materials directly targeting PA knowledge and skills was considerably lower. Specifically, approximately half of the participants viewed online lectures about PA (54.7%), PA promotion posters (46.7%), and online lectures about exercise (46.0%). This gap suggests that although participants maintained their participation in the intervention, they did not engage deeply enough with the core educational content required to modify their PA behaviours.

This study had several strengths. First, to the best of our knowledge, this is one of the first studies to use a rigorous cluster randomised controlled trial design to evaluate an occupational lifestyle intervention specifically targeting teleworkers. This design enabled us to account for workplace-level effects and minimise contamination risk. Second, the outcome measures were detailed and objective, which allowed stratification of results by activity type and intensity (ST, LPA, and MVPA). Furthermore, by combining these objective data with a daily diary (capturing the work context), we conducted subdomain analyses. These provided a detailed understanding of PA patterns across workdays, weekends, teleworking days, and commuting days. Third, the intervention was designed to be low-cost and highly scalable. By using remote delivery without incurring high costs or imposing financial burdens on participating companies, we provide valuable data on the effectiveness of a pragmatic and feasible public health approach.

However, this study had some limitations. First, this study was probably underpowered. The final recruited sample (n=310) did not meet the target sample size (n=500) due to the length of the research grant contract. Second, the interpretation of our null finding was difficult due to population heterogeneity and potentially uncontrolled confounding variables. Changes in teleworking frequency before and after the intervention may have acted as a significant confounder, diluting any actual intervention effect, although we could not assess these changes. Finally, our findings may not be generalisable because we relied on a non-probability snowball approach with individual volunteers, predominantly from the Tokyo metropolitan area.

In conclusion, this 12-week multicomponent intervention did not significantly increase daily step count or improve other PA outcomes among Japanese teleworkers. Suboptimal intervention implementation and a significant loss of statistical power associated with recruitment constraints likely explain this result. These findings suggest that future research should diversify intervention strategies and apply more precise eligibility criteria to target heterogeneous teleworking populations effectively.

## Supporting information

CONSORT 2025 statement checklist

CONSORT 2010 statement: extension to cluster randomised trials checklist

Telework_supplementary_table_1

## Data Availability

All data produced in the present study are available upon reasonable request to the authors

## Contributors

J.K. drafted the manuscript. J.K. and Y.N. directly accessed and verified the underlying data reported in this manuscript. M.G. contributed to the statistical analysis. A.W., S.K., T.Y., R.T., W.U., T.S., N.Y., K.Y., M.G., and Y.K. contributed to the study conception, protocol development, and critical revision of the manuscript. All authors approved the final version of the manuscript.

## Data Sharing

De-identified participant-level data are available from the corresponding author upon reasonable request.

## Acknowledgements

We sincerely thank all participants in the TELEWORK study, as well as the six participating companies and their workplace coordinators, for their invaluable collaboration and support during the implementation phase.

## References

1. Athanasiadou C, Theriou G. Telework: systematic literature review and future research agenda. Heliyon 2021; 7: e08165.

2. Ministry of Land, Infrastructure, Transport and Tourism. FY2024 Survey on the Actual Status of the Teleworking Population – Survey Results. Accessed October 1, Japanese, 2025, https://www.mlit.go.jp/toshi/kankyo/telework_index.html

3. Fukushima N, Machida M, Kikuchi H, et al. Associations of working from home with occupational physical activity and sedentary behavior under the COVID-19 pandemic. J Occup Health 2021; 63: e12212.

4. Kitano N, Fujii Y, Wada A, et al. Associations of working from home frequency with accelerometer-measured physical activity and sedentary behavior in japanese white-collar workers: a cross-sectional analysis of the Meiji Yasuda LifeStyle Study. J Phys Act Health 2024; 21: 1150–7.

5. Garcia L, Pearce M, Abbas A, et al. Non-occupational physical activity and risk of cardiovascular disease, cancer and mortality outcomes: a dose-response meta-analysis of large prospective studies. Br J Sports Med 2023; 57: 979–89.

6. Patterson R, McNamara E, Tainio M, et al. Sedentary behaviour and risk of all-cause, cardiovascular and cancer mortality, and incident type 2 diabetes: a systematic review and dose response meta-analysis. Eur J Epidemiol 2018; 33: 811–29.

7. Wütschert MS, Romano-Pereira D, Suter L, Schulze H, Elfering A. A systematic review of working conditions and occupational health in home office. Work 2022; 72: 839–52.

8. Kanamori S, Tabuchi T, Kai Y. Association between the telecommuting environment and somatic symptoms among teleworkers in Japan. J Occup Health 2024; 66: uiad014.

9. Watanabe K, Kawakami N. Effects of a multi-component workplace intervention program with environmental changes on physical activity among japanese white-collar employees: a cluster-randomized controlled trial. Int J Behav Med 2018; 25: 637–48.

10. Healy GN, Eakin EG, Owen N, et al. A cluster randomized controlled trial to reduce office workers’ sitting time: effect on activity outcomes. Med Sci Sports Exerc 2016; 48: 1787–97.

11. Edwardson CL, Yates T, Biddle SJH, et al. Effectiveness of the Stand More AT (SMArT) Work intervention: cluster randomised controlled trial. BMJ 2018; 363: k3870.

12. Edwardson CL, Biddle SJH, Clemes SA, et al. Effectiveness of an intervention for reducing sitting time and improving health in office workers: three arm cluster randomised controlled trial. BMJ 2022; 378: e069288.

13. Brakenridge CL, Fjeldsoe BS, Young DC, et al. Evaluating the effectiveness of organisational-level strategies with or without an activity tracker to reduce office workers’ sitting time: a cluster-randomised trial. Int J Behav Nutr Phys Act 2016; 13: 115.

14. Lyzwinski LN. Organizational and occupational health issues with working remotely during the pandemic: a scoping review of remote work and health. J Occup Health 2024; 66: uiae005.

15. Peristera P, Bergljottsdotter C, Leineweber C. When home becomes the office: navigating challenges and embracing possibilities in telework in Sweden during and after the COVID-19 pandemic. Front Psychol 2025; 16: 1516074.

16. Wada A, Kim J, Kanamori S, et al. Multicomponent occupational lifestyle intervention to improve physical activity, musculoskeletal health, and work environment among Japanese teleworkers (TELEWORK study): protocol for a cluster randomized controlled trial. J Occup Health 2025; 67: uiaf014.

17. Torgerson DJ. Contamination in trials: is cluster randomisation the answer? BMJ 2001; 322: 355–7

18. Kim J, Mizushima R, Morimoto M, et al. Effectiveness of a multicomponent intervention to promote physical activity among Japanese remote workers: a pilot open-label randomized controlled trial. J Occup Health 2024; 66: uiae052.

19. Hopewell S, Chan AW, Collins GS, et al. CONSORT 2025 statement: updated guideline for reporting randomised trials. Lancet. Published online April 14, 2025.

20. Campbell MK, Piaggio G, Elbourne DR, Altman DG; CONSORT Group. Consort 2010 statement: extension to cluster randomised trials. BMJ 2012; 345: e5661.

21. Sallis JF, Owen N, Fisher EB. Ecological models of health behavior. In: Glanz K, Rimer BK, Viswanath K, editors. Health Behavior and Health Education: Theory, Research, and Practice. 4th ed. New York: Jossey-Bass A Wiley Imprint; 2008.

22. Miyachi M, Tripette J, Kawakami R, Murakami H. "+10 min of physical activity per day": Japan is looking for efficient but feasible recommendations for its population. J Nutr Sci Vitaminol (Tokyo) 2015; 61: S7–9.

23. Owen N, Healy GN, Matthews CE, Dunstan DW. Too much sitting: the population health science of sedentary behavior. Exerc Sport Sci Rev 2010; 38: 105–13.

24. Diaz KM, Howard VJ, Hutto B, et al. Patterns of sedentary behavior and mortality in U.S. middle-aged and older adults: a national cohort study. Ann Intern Med 2017; 167: 465–75.

25. Yoshinaga S, Shiomitsu T, Kamohara M, Fujii Y, Chosa E, Tsuruta K. Lifestyle-related signs of locomotive syndrome in the general Japanese population: a cross-sectional study. J Orthop Sci 2019; 24: 1105–9.

26. Review Committee on the Revision of Physical Activity Reference for Health Promotion 2013. Physical Activity Guide for Health Promotion 2023. Health Service Bureau, Ministry of Health, Labour and Welfare; 2024.

27. Bull FC, Al-Ansari SS, Biddle S, et al. World Health Organization 2020 guidelines on physical activity and sedentary behaviour. Br J Sports Med 2020; 54: 1451–62.

28. Ohkawara K, Oshima Y, Hikihara Y, Ishikawa-Takata K, Tabata I, Tanaka S. Real-time estimation of daily physical activity intensity by a triaxial accelerometer and a gravity-removal classification algorithm. Br J Nutr 2011; 105: 1681–91.

29. Oshima Y, Kawaguchi K, Tanaka S, et al. Classifying household and locomotive activities using a triaxial accelerometer. Gait Posture 2010; 31: 370–4.

30. Troiano RP, Berrigan D, Dodd KW, Mâsse LC, Tilert T, McDowell M. Physical activity in the United States measured by accelerometer. Med Sci Sports Exerc 2008; 40: 181–8.

31. Banach M, Lewek J, Surma S, et al. The association between daily step count and all-cause and cardiovascular mortality: a meta-analysis. Eur J Prev Cardiol 2023; 30: 1975–85.

32. Barnard J, Rubin DB. Small-sample degrees of freedom with multiple imputation. Biometrika 1999; 86: 948–55.

33. Huang S, Fiero MH, Bell ML. Generalized estimating equations in cluster randomized trials with a small number of clusters: review of practice and simulation study. Clin Trials 2016; 13: 445–9.

34. Pustejovsky JE, Tipton E. Small-sample methods for cluster-robust variance estimation and hypothesis testing in fixed effects models. J Bus Econ Stat 2018; 36: 672–83.

35. Jirathananuwat A, Pongpirul K. Promoting physical activity in the workplace: a systematic meta-review. J Occup Health 2017; 59: 385–93.

36. Kramer JN, Tinschert P, Scholz U, Fleisch E, Kowatsch T. A Cluster-randomized trial on small incentives to promote physical activity. Am J Prev Med 2019; 56: e45–54.

37. Patel MS, Small DS, Harrison JD, et al. Effectiveness of behaviorally designed gamification interventions with social incentives for increasing physical activity among overweight and obese adults across the United States: The STEP UP randomized clinical trial. JAMA Intern Med 2019; 179: 1624–32.

38. Kim J, Mizushima R, Nishida K, Morimoto M, Nakata Y. Proposal of a comprehensive and multi-component approach to promote physical activity among Japanese office workers: a qualitative focus group interview study. Int J Environ Res Public Health 2022a; 19: 2172.

39. Munir F, Biddle SJH, Davies MJ, et al. Stand More AT Work (SMArT Work): using the behaviour change wheel to develop an intervention to reduce sitting time in the workplace. BMC Public Health 2018; 18: 319.

