## Supplementary material for "Effectiveness of a 12-week multicomponent occupational lifestyle intervention to increase physical activity among Japanese teleworkers: a cluster randomised controlled trial (TELEWORK study)": CONSORT 2025 statement checklist

The CONSORT reporting checklist

For checking that reports of randomised trials can be understood and used by everyone

How to use this reporting checklist

This reporting checklist allows authors to demonstrate that their manuscripts adhere to the [CONSORT reporting guideline](https:/resources.equator-network.org/reporting-guidelines/consort/index.html).

If you have not used a reporting guideline before, read about [how and why to use them](https:/resources.equator-network.org/about/reporting-guidelines.html) and check whether CONSORT is the [most applicable reporting guideline](https:/resources.equator-network.org/reporting-guidelines/consort/index.html?#applicability) for your work.

Reporting guidelines are most useful when used early in research. When writing a manuscript or application, consider using the [full guidance](https:/resources.equator-network.org/reporting-guidelines/consort/index.html) where you’ll find explanations and examples for each item.

After writing, demonstrate adherence by completing this checklist:

1. Specify where each item is described (see Note 1).
2. Cite this checklist (See Note 2).
3. Include your completed checklist as a supplement when submitting to a journal so that future readers can use it to find information.

|  | Item Description | Location (or reason for not reporting) |
| --- | --- | --- |
| **Title and Abstract** |  |  |
| [1a. Title](https:/resources.equator-network.org/reporting-guidelines/consort/items/title.html?utm_source=consort&utm_medium=checklist&utm_campaign=CONSORT_2025_v1_1) | Identification as a randomised trial. | Page 1 |
| [1b. Structured Abstract](https:/resources.equator-network.org/reporting-guidelines/consort/items/structured-abstract.html?utm_source=consort&utm_medium=checklist&utm_campaign=CONSORT_2025_v1_1) | Structured summary of the trial design, methods, results, and conclusions. | Page 3-4 |
| **Open Science** |  |  |
| [2. Trial Registration](https:/resources.equator-network.org/reporting-guidelines/consort/items/trial-registration.html?utm_source=consort&utm_medium=checklist&utm_campaign=CONSORT_2025_v1_1) | Name of trial registry, identifying number (with URL) and date of registration. | Page 21  <https://center6.umin.ac.jp/cgi-open-bin/ctr_e/ctr_view.cgi?recptno=R000061478> |
| [Protocol and statistical analysis plan](https:/resources.equator-network.org/reporting-guidelines/consort/items/protocol-and-statistical-analysis-plan.html?utm_source=consort&utm_medium=checklist&utm_campaign=CONSORT_2025_v1_1) | Where the trial protocol and statistical analysis plan can be accessed. | Page 21  doi: 10.1093/joccuh/uiaf014 |
| [4. Data sharing](https:/resources.equator-network.org/reporting-guidelines/consort/items/data-sharing.html?utm_source=consort&utm_medium=checklist&utm_campaign=CONSORT_2025_v1_1) | Where and how the individual de-identified participant data (including data dictionary), statistical code and any other materials can be accessed. | Page 31 |
| 5. Funding and Conflicts of Interest |  |  |
| [5a. Funding](https:/resources.equator-network.org/reporting-guidelines/consort/items/funding.html?utm_source=consort&utm_medium=checklist&utm_campaign=CONSORT_2025_v1_1) | Sources of funding and other support (eg, supply of drugs), and role of funders in the design, conduct, analysis, and reporting of the trial. | Page 31 |
| [5b. Conflicts of interest](https:/resources.equator-network.org/reporting-guidelines/consort/items/conflicts-of-interest.html?utm_source=consort&utm_medium=checklist&utm_campaign=CONSORT_2025_v1_1) | Financial and other conflicts of interest of the manuscript authors. | Page 31 |
| **Introduction** |  |  |
| [6. Background and rationale](https:/resources.equator-network.org/reporting-guidelines/consort/items/background-and-rationale.html?utm_source=consort&utm_medium=checklist&utm_campaign=CONSORT_2025_v1_1) | Scientific background and rationale. | Page 8-9 |
| [7. Objectives](https:/resources.equator-network.org/reporting-guidelines/consort/items/objectives.html?utm_source=consort&utm_medium=checklist&utm_campaign=CONSORT_2025_v1_1) | Specific objectives related to benefits and harms. | Page 8-9 |
| **Methods** |  |  |
| [8. Patient and public involvement](https:/resources.equator-network.org/reporting-guidelines/consort/items/patient-and-public-involvement.html?utm_source=consort&utm_medium=checklist&utm_campaign=CONSORT_2025_v1_1) | Details of patient or public involvement in the design, conduct and reporting of the trial. | N/A |
| [9. Trial Design](https:/resources.equator-network.org/reporting-guidelines/consort/items/trial-design.html?utm_source=consort&utm_medium=checklist&utm_campaign=CONSORT_2025_v1_1) | Description of trial design including type of trial (eg, parallel group, crossover), allocation ratio, and framework (eg, superiority, equivalence, non-inferiority, exploratory). | Page 10 |
| [10. Changes to trial protocol](https:/resources.equator-network.org/reporting-guidelines/consort/items/changes-to-trial-protocol.html?utm_source=consort&utm_medium=checklist&utm_campaign=CONSORT_2025_v1_1) | Important changes to the trial after it commenced including any outcomes or analyses that were not pre-specified, with reason. | Page 20 |
| [11. Trial Setting](https:/resources.equator-network.org/reporting-guidelines/consort/items/trial-setting.html?utm_source=consort&utm_medium=checklist&utm_campaign=CONSORT_2025_v1_1) | Settings (eg, community, hospital) and locations (eg, countries, sites) where the trial was conducted. | Page 10 |
| 12. Eligibility Criteria |  |  |
| [12a. Participants](https:/resources.equator-network.org/reporting-guidelines/consort/items/eligibility-criteria-participants.html?utm_source=consort&utm_medium=checklist&utm_campaign=CONSORT_2025_v1_1) | Eligibility criteria for participants. | Page 10-11 |
| [12b. Other](https:/resources.equator-network.org/reporting-guidelines/consort/items/eligibility-criteria-sites-individuals.html?utm_source=consort&utm_medium=checklist&utm_campaign=CONSORT_2025_v1_1) | If applicable, eligibility criteria for sites and for individuals delivering the interventions (eg, surgeons, physiotherapists). | N/A |
| [13. Intervention and comparator](https:/resources.equator-network.org/reporting-guidelines/consort/items/intervention-and-comparator.html?utm_source=consort&utm_medium=checklist&utm_campaign=CONSORT_2025_v1_1) | Intervention and comparator with sufficient details to allow replication. If relevant, where additional materials describing the intervention and comparator (eg, intervention manual) can be accessed. | Page 12-16 |
| [14. Outcomes](https:/resources.equator-network.org/reporting-guidelines/consort/items/outcomes.html?utm_source=consort&utm_medium=checklist&utm_campaign=CONSORT_2025_v1_1) | Prespecified primary and secondary outcomes, including the specific measurement variable (eg, systolic blood pressure), analysis metric (eg, change from baseline, final value, time to event), method of aggregation (eg, median, proportion), and time point for each outcome. | Page 16-18 |
| [15. Harms](https:/resources.equator-network.org/reporting-guidelines/consort/items/harms.html?utm_source=consort&utm_medium=checklist&utm_campaign=CONSORT_2025_v1_1) | How harms were defined and assessed (eg, systematically, non-systematically). | Page 18 |
| 16. Sample Size |  |  |
| [16a. How sample size was determined](https:/resources.equator-network.org/reporting-guidelines/consort/items/sample-size-determination.html?utm_source=consort&utm_medium=checklist&utm_campaign=CONSORT_2025_v1_1) | How sample size was determined, including all assumptions supporting the sample size calculation. | Page 18 |
| [16b. Interim analyses and stopping criteria](https:/resources.equator-network.org/reporting-guidelines/consort/items/sample-size-interim-analyses-and-stopping-guidelines.html?utm_source=consort&utm_medium=checklist&utm_campaign=CONSORT_2025_v1_1) | Explanation of any interim analyses and stopping guidelines. | N/A |
| 17. Randomisation |  |  |
| [17a. Sequence Generation](https:/resources.equator-network.org/reporting-guidelines/consort/items/randomisation-sequence-generation.html?utm_source=consort&utm_medium=checklist&utm_campaign=CONSORT_2025_v1_1) | Who generated the random allocation sequence and the method used. | Page 11-12 |
| [17b. Type of Randomisation](https:/resources.equator-network.org/reporting-guidelines/consort/items/randomisation-type-of-randomisation.html?utm_source=consort&utm_medium=checklist&utm_campaign=CONSORT_2025_v1_1) | Type of randomisation and details of any restriction (eg, stratification, blocking, and block size). | Page 11-12 |
| [18. Allocation concealment mechanism](https:/resources.equator-network.org/reporting-guidelines/consort/items/allocation-concealment-mechanism.html?utm_source=consort&utm_medium=checklist&utm_campaign=CONSORT_2025_v1_1) | Mechanism used to implement the random allocation sequence (eg, central computer/telephone; sequentially numbered, opaque, sealed containers), describing any steps to conceal the sequence until interventions were assigned. | Page 11-12 |
| [19. Implementation](https:/resources.equator-network.org/reporting-guidelines/consort/items/implementation.html?utm_source=consort&utm_medium=checklist&utm_campaign=CONSORT_2025_v1_1) | Whether the personnel who enrolled and those who assigned participants to the interventions had access to the random allocation sequence. | Page 11-12 |
| 20. Blinding |  |  |
| [20a. Who was blinded](https:/resources.equator-network.org/reporting-guidelines/consort/items/blinding-who.html?utm_source=consort&utm_medium=checklist&utm_campaign=CONSORT_2025_v1_1) | Who was blinded after assignment to interventions (eg, participants, care providers, outcome assessors, data analysts). | Page 11-12 |
| [20b. How blinding was achieved](https:/resources.equator-network.org/reporting-guidelines/consort/items/blinding-how.html?utm_source=consort&utm_medium=checklist&utm_campaign=CONSORT_2025_v1_1) | If blinded, how blinding was achieved and description of the similarity of interventions. | N/A |
| 21. Statistical methods |  |  |
| [21a. Comparing groups](https:/resources.equator-network.org/reporting-guidelines/consort/items/statistical-methods-comparing-groups-primary-secondary-outcomes-harms.html?utm_source=consort&utm_medium=checklist&utm_campaign=CONSORT_2025_v1_1) | Statistical methods used to compare groups for primary and secondary outcomes, including harms. | Page 19-21 |
| [21b. Definition of who is included in each analysis](https:/resources.equator-network.org/reporting-guidelines/consort/items/statistical-methods-definition-of-who-is-included-in-each-analysis.html?utm_source=consort&utm_medium=checklist&utm_campaign=CONSORT_2025_v1_1) | Definition of who is included in each analysis (e.g., all randomised participants), and in which group. | Page 19-21 |
| [21c. Missing Data](https:/resources.equator-network.org/reporting-guidelines/consort/items/statistical-methods-missing-data.html?utm_source=consort&utm_medium=checklist&utm_campaign=CONSORT_2025_v1_1) | How missing data were handled in the analysis. | Page 19-21 |
| [21d. Additional Analyses](https:/resources.equator-network.org/reporting-guidelines/consort/items/statistical-methods-additional-analyses.html?utm_source=consort&utm_medium=checklist&utm_campaign=CONSORT_2025_v1_1) | Methods for any additional analyses (eg, subgroup and sensitivity analyses), distinguishing pre-specified from post hoc. | Page 19-21 |
| 22. Participant flow, including flow diagram |  |  |
| [22a. Participant Numbers](https:/resources.equator-network.org/reporting-guidelines/consort/items/participant-flow-numbers.html?utm_source=consort&utm_medium=checklist&utm_campaign=CONSORT_2025_v1_1) | For each group, the numbers of participants who were randomly assigned, received intended intervention, and were analysed for the primary outcome. | Page 21-22, Figure 1 |
| [22b. Losses and exclusions](https:/resources.equator-network.org/reporting-guidelines/consort/items/participant-flow-losses-and-exclusions.html?utm_source=consort&utm_medium=checklist&utm_campaign=CONSORT_2025_v1_1) | For each group, losses and exclusions after randomisation, together with reasons. | Page 21-22, Figure 1 |
| 23. Recruitment |  |  |
| [23a. Dates](https:/resources.equator-network.org/reporting-guidelines/consort/items/recruitment-dates.html?utm_source=consort&utm_medium=checklist&utm_campaign=CONSORT_2025_v1_1) | Dates defining the periods of recruitment and follow-up for outcomes of benefits and harms. | Page 10 |
| [23b. Reasons for stopping recruitment](https:/resources.equator-network.org/reporting-guidelines/consort/items/recruitment-why-stopped.html?utm_source=consort&utm_medium=checklist&utm_campaign=CONSORT_2025_v1_1) | If relevant, why the trial ended or was stopped. | N/A |
| 24. Intervention and comparator delivery |  |  |
| [24a. As Administered](https:/resources.equator-network.org/reporting-guidelines/consort/items/intervention-comparator-delivery-as-administered.html?utm_source=consort&utm_medium=checklist&utm_campaign=CONSORT_2025_v1_1) | Intervention and comparator as they were actually administered (eg, where appropriate, who delivered the intervention/comparator, whether participants adhered, whether they were delivered as intended (fidelity)). | Page 22 |
| [24b. Concomitant Care](https:/resources.equator-network.org/reporting-guidelines/consort/items/intervention-comparator-delivery-concomitant-care.html?utm_source=consort&utm_medium=checklist&utm_campaign=CONSORT_2025_v1_1) | Concomitant care received during the trial for each group. | Page 16 |
| [25. Baseline Data](https:/resources.equator-network.org/reporting-guidelines/consort/items/baseline-data.html?utm_source=consort&utm_medium=checklist&utm_campaign=CONSORT_2025_v1_1) | A table showing baseline demographic and clinical characteristics for each group. | Page 22-23 |
| [26. Numbers analysed, outcomes, and estimation](https:/resources.equator-network.org/reporting-guidelines/consort/items/numbers-analysed-outcomes-estimation.html?utm_source=consort&utm_medium=checklist&utm_campaign=CONSORT_2025_v1_1) | For each primary and secondary outcome, by group:   - the number of participants included in the analysis. - the number of participants with available data at the outcome time point. - result for each group, and the estimated effect size and its precision (such as 95% confidence interval). - for binary outcomes, presentation of both absolute and relative effect size. | Page 21-25, 41-51 |
| [27. Harms](https:/resources.equator-network.org/reporting-guidelines/consort/items/harms.html?utm_source=consort&utm_medium=checklist&utm_campaign=CONSORT_2025_v1_1) | All harms or unintended events in each group. | Page 25-26 |
| [28. Ancillary Analyses](https:/resources.equator-network.org/reporting-guidelines/consort/items/ancillary-analyses.html?utm_source=consort&utm_medium=checklist&utm_campaign=CONSORT_2025_v1_1) | Any other analyses performed, including subgroup and sensitivity analyses, distinguishing pre-specified from post hoc. | Page 24-25 |
| **Discussion** |  |  |
| [29. Interpretation](https:/resources.equator-network.org/reporting-guidelines/consort/items/interpretation.html?utm_source=consort&utm_medium=checklist&utm_campaign=CONSORT_2025_v1_1) | Interpretation consistent with results, balancing benefits and harms, and considering other relevant evidence. | Page 26-30 |
| [30. Limitations](https:/resources.equator-network.org/reporting-guidelines/consort/items/limitations.html?utm_source=consort&utm_medium=checklist&utm_campaign=CONSORT_2025_v1_1) | Trial limitations, addressing sources of potential bias, imprecision, generalisability, and, if relevant, multiplicity of analyses. | Page 26-30 |

### 1 How to specify where content is

Tell the reader where they can find information. E.g.,

- Results; paragraph 2
- Methods, Participants; paragraphs 1 & 2.
- Table 3
- Supplement B, para. 4

If you have chosen not to describe an item, explain why. You can do this in the checklist, or as a note below it.

You can describe items in the article body, or in tables, figures, or supplementary materials, and should prioritize items you feel are most important to your intended audience. The order of items in your manuscript does not need to match the order of items in this checklist. You can decide how best to structure your work.

### 2 How to cite

Describe how you used CONSORT at the end of your Methods section, referencing the resources you used e.g.,

‘We used the CONSORT reporting guideline(1) to draft this manuscript, and the CONSORT reporting checklist(2) when editing, included in supplement A’

If you use a reporting checklist, remember to include it as a supplement when publishing so that readers can easily find information and see how you have interpreted the guidance.

1.	Hopewell S, Chan AW, Collins GS, Hróbjartsson A, Moher D, Schulz KF, et al. CONSORT 2025 statement: Updated guideline for reporting randomised trials. The BMJ [Internet]. 2025 Apr;389:e081123. Available from: <https://www.ncbi.nlm.nih.gov/pmc/articles/PMC11995449/>

2.	Hopewell S, Chan AW, Collins GS, Hróbjartsson A, Moher D, Schulz KF, et al. The CONSORT reporting checklist. In: Harwood J, Albury C, Beyer J de, Schlüssel M, Collins G, editors. The EQUATOR network reporting guideline platform [Internet]. The UK EQUATOR Centre; 2025. Available from: [https:/resources.equator-network.org/reporting-guidelines/consort/consort-checklist.docx](https://https:/resources.equator-network.org/reporting-guidelines/consort/consort-checklist.docx)
