## Supplementary material for "Effectiveness of a 12-week multicomponent occupational lifestyle intervention to increase physical activity among Japanese teleworkers: a cluster randomised controlled trial (TELEWORK study)": CONSORT 2010 statement: extension to cluster randomised trials checklist

**Cluster RCT Extension Checklist**

*Campbell MK et al. CONSORT 2010 statement: extension to cluster randomised trials. BMJ 2012;345:e5661*

| **Item No.** | **Cluster RCT Extension Item** | **Location (or reason for not reporting)** |
| --- | --- | --- |
| **TITLE AND ABSTRACT** | | |
| **1a** | Identify the study specifically as a cluster randomised trial in the title. | Page 1 |
| **1b** | In the abstract, additionally report: • Eligibility criteria for clusters • How clusters were allocated to interventions • Number of clusters randomised and analysed per group • Primary outcome results at cluster or individual participant level as applicable | Page 3–4 |
| **INTRODUCTION** | | |
| **2a (→Item 6)** | Provide rationale for using a cluster design (eg, intervention delivered at cluster level, contamination prevention, administrative convenience). | Page 8–9 |
| **2b (→Item 7)** | State whether objectives pertain to the cluster level, the individual participant level, or both. | Page 8–9 |
| **METHODS** | | |
| **3a (→Item 9)** | Define what constitutes a cluster. Describe how the design features (allocation, intervention, analysis) apply at the cluster level. | Page 10 |
| **4a (→Item 12a)** | State eligibility criteria for clusters as well as for individual participants. | Page 10–11 |
| **5 (→Item 13)** | State whether interventions are delivered at the cluster level, the individual participant level, or both. | Page 12–16 |
| **6a (→Item 14)** | State whether outcome measures pertain to the cluster level, the individual participant level, or both. | Page 16–18 |
| **7a (→Item 16a)** | Report method of sample size calculation specifying: • Number of clusters (and whether equal or unequal cluster sizes assumed) • Assumed cluster size • Intracluster correlation coefficient (ICC or k) and its uncertainty/source | Page 18 |
| **8b (→Item 17b)** | Provide details of stratification or matching at the cluster level, if used. | Page 11–12 |
| **9 (→Item 18)** | Specify that allocation was based on clusters rather than individuals. State whether allocation concealment was at the cluster level, individual level, or both. | Page 11–12 |
| **10a (→Item 19)** | Who generated the allocation sequence, who enrolled clusters, and who assigned clusters to interventions. | Page 11–12 |
| **10b (→Item 19)** | Mechanism by which individual participants were included in clusters for the trial (eg, complete enumeration, random sampling). | Page 11–12 |
| **10c (→Item 19)** | From whom consent was sought (cluster representatives, individual members, or both), and whether consent was obtained before or after randomisation. | Page 11–12 |
| **12a (→Item 21a)** | Describe explicitly how clustering was taken into account in the statistical analysis (eg, mixed-effects models, GEE, multilevel models). | Page 19–21 |
| **RESULTS** | | |
| **13a (→Item 22a)** | For each group, report the number of clusters randomly assigned, that received the intended treatment, and that were analysed for the primary outcome. | Page 21–22, Figure 1 |
| **13b (→Item 22b)** | Report losses and exclusions separately for clusters and for individual cluster members, with reasons. | Page 21–22, Figure 1 |
| **15 (→Item 25)** | Present baseline characteristics at both the individual and cluster level for each group, as applicable. | Page 22–23, Table 1 |
| **16 (→Item 26)** | Report the number of clusters included in each analysis per group. | Page 21–25 |
| **17a (→Item 26)** | Report results at the individual or cluster level as applicable. Report the intracluster correlation coefficient (ICC or k) for each primary outcome. | Page 21–25, Table 2 |
| **DISCUSSION** | | |
| **21 (→Item 30)** | Address generalisability separately to clusters and to individual participants, as relevant. | Page 26–30 |

** Location: record paragraph, section, table, figure, or supplement where the item is reported. If not reported, state reason.*
