## Supplementary material for "Effectiveness of a 12-week multicomponent occupational lifestyle intervention to increase physical activity among Japanese teleworkers: a cluster randomised controlled trial (TELEWORK study)": Telework_supplementary_table_1

**Supplementary Table**
**Supplementary Table 1. Changes in physical activity outcomes from baseline to week 12: GEE analysis without multiple imputations in the FAS.**

|  | **Intervention, n=156  Mean (SD)** | **Control, n=154 Mean (SD)** | **Adjusted mean difference (Intervention − control)  95% CI** | **† p-value** | **ICC** |
| --- | --- | --- | --- | --- | --- |
| **Primary outcome** |  |  |  |  |  |
| Daily steps, steps/day |  |  |  |  |  |
| Baseline | 7,036 (2,803) | 6,941 (2,971) |  |  |  |
| Week 12 | 7,280 (3,440) | 6,931 (2,872) |  |  |  |
| Change from baseline | 184 (2,432) | −49 (2,133) | 280 (−382, 942) | 0·364 | 0·043 |
| **Secondary outcome** |  |  |  |  |  |
| Sedentary time, min/day |  |  |  |  |  |
| Baseline | 638·0 (106·8) | 633·4 (108·1) |  |  |  |
| Week 12 | 630·3 (114·1) | 635·4 (107·3) |  |  |  |
| Change from baseline | −10·6 (74·5) | 0·7 (77·2) | −4·4 (−17·5, 8·6) | 0·464 | 0·029 |
| Light-intensity physical activity, min/day |  |  |  |  |  |
| Baseline | 185·8 (54·6) | 177·1 (67·2) |  |  |  |
| Week 12 | 182·9 (60·9) | 175·3 (71·1) |  |  |  |
| Change from baseline | −5·8 (49·3) | −2·5 (39·7) | 0·8 (−10·9, 12·6) | 0·879 | 0·068 |
| Moderate to vigorous intensity physical activity, min/day |  |  |  |  |  |
| Baseline | 54·2 (24·2) | 54·1 (24·9) |  |  |  |
| Week 12 | 57·6 (28·1) | 53·9 (23·7) |  |  |  |
| Change from baseline | 2·6 (19·7) | 0·5 (16·7) | 3·6 (−1·6, 8·8) | 0·149 | 0·039 |

* Values are presented as mean (SD) unless otherwise indicated. Analyses were conducted on the full analysis set, which included all participants who were randomised according to the intention-to-treat principle. No multiple imputations were performed. Within-group changes and adjusted mean differences were estimated using generalised estimating equations (GEE) with an exchangeable correlation structure that accounted for clustering at the worksite level. The model was adjusted for the accelerometer wear time.

† *p*-value for the group × time interaction term. CI, confidence interval; FAS, full analysis set; ICC, intraclass correlation coefficient; SD, standard deviation..
